# Government restriction efficiency on curbing COVID-19 pandemic transmission in Western Europe

**DOI:** 10.1101/2022.02.28.22271636

**Authors:** Simone Lolli, Gemine Vivone

**Affiliations:** CNR-IMAA, Contrada S. Loja, 85050 Tito Scalo (PZ), Italy

## Abstract

In this article we assess the effectiveness of the restrictions, implemented at government level, to curb the first wave of the COVID-19 outbreak (January-May 2020) in seven European countries. The analysis put in evidence a strong correlation (correlation coefficient greater than 0.85) between one of the statistical parameters of the distribution representing the temporal evolution of the weekly number of patients admitted into the Intensive Cure Unit (ICU), i.e., the skewness, and the Stringency Index, an aggregate synthetic variable that reflects the level of implemented restrictions by a specific country on a scale between 0 (no restrictions) and 100 (full lockdown). Then, to assess if the skewness is consistent in effectively reflecting the applied restrictions, we computed the skewness for non-Covid flu outbreaks during four years from 2014-2015 to 2018-2019 and Covid-19 outbreak, where no restrictions were applied in Italy. The results highlight a significant difference between the values of the skewness for the normal flu with respect to the COVID-19 outbreak. This large difference put in evidence that the implemented restrictions modify the skewness of the ICU hospitalized patient number distribution. The skewness then can be used as a valid indicator to assess if the restriction had any effect on pandemic transmission and can be used as a support for decision makers.

## Introduction

The World Health Organization (WHO) confirmed pandemic (March 2020) the new Severe Acute Respiratory Syndrome Coronavirus 2 (SARS-CoV-2) infection that originated in Wuhan, China, in December 2019 (first reported cases), then spreading to Italy and successively worldwide. Some recent studies highlight how the meteorological conditions and air-pollution influenced the pandemic transmission in several parts of the world [1–7] other than the social and economical factors [8, 9] or how fast asymptomatic carriers are found [10]. However, to curb the outbreak transmission, following the Chinese example, the European governments implemented different and progressive restrictions, as imposing face masks indoor and/or outdoor, closing schools, restaurants, theaters, canceling sport events, concerts and any social activity gathering public, up to ban people from traveling and leaving their homes except for unavoidable and urgent needs, e.g., grocery, medical visits [11]. The applied restrictions were not uniformly applied in Europe. They also produced a considerable negative impact on the economy [12]. In this article, we assess, from a statistical point of view, the effectiveness of the applied restrictions in seven European western countries as Italy, France, Sweden, Finland, Germany, Belgium and The Netherlands. Other studies tried to assess the restriction effectiveness [11], but using indexes or variables strongly dependent on testing policy or using models [13]. In this analysis, for the first time, we assess a possible correlation between the different statistical parameters characterizing the distribution of the weekly number of patients hospitalized into the Intensive Care Unit (ICU) in the seven European countries and the averaged Stringency Index (SI; [15]), a synthetic index that represents a metric to quantify the severity of the applied restriction. It is represented by a single number obtained aggregating the different restriction indexes and it ranges from 0 (no restriction) to 100 (full lockdown). Once assessing which statistical parameter of the ICU distribution shows the stronger correlation with the SI, we test its significance level, i.e. if the statistical parameter is consistent and then it is able to effectively be a proxy of the implemented restrictions. To validate the latter speculation, we check if there is a significant difference of the considered statistical parameter values for COVID-19 and the distribution of ICU hospitalized patients during several pre-pandemic flu outbreaks where no restrictions were applied in Italy (data not available for the other countries).

We chose, among all the possible variables, the hospitalized ICU patient number because this variable is much more reliable with respect to other ones, e.g., the weekly number of people tested positive [14]. The hospitalized ICU patient number is then a proxy to assess the effective infection rate.

## Methodology

The objective of this study is to assess the restriction effectiveness from a statistical point of view. To reach this goal, we quantitatively evaluate the differences in the evolution of the number of patients hospitalized in the ICU over time for seven western European nations who applied different levels of restrictions that can be summarized with the Stringency Index (SI; [15]). This index provides a number ranging from 0 to 100 that reflects the level of implemented restrictions by a certain country (0 no restrictions, 100 full lockdown, all the non-vital activities are suspended). The SI index is based on the aggregation of eight closure policies and containment indicators and one public information campaign indicator. Each imposed restriction leads to an increment of the SI for the related country. The eight closure policies and containment indicators are listed in Tab.1.

**Table 1.** Main indicators contributing to the Stringency Index value: 0 no restrictions, 100 full lockdown.

| Indicator |
| --- |
| School Closing |
| Workplace Closing |
| Canceling Public Events |
| Restriction on gathering size |
| Close Public Transport |
| Stay at Home Requirements |
| Restrictions on Internal Movements |
| Restrictions on International Travels |

The higher the SI, the greater the number of implemented restrictions. Data are freely available from the University of Oxford website. The SI values are available for each country on daily basis, but, for our purposes and to avoid the volatility of the index, we simply averaged the values along the first pandemic wave (January-May 2020).

By the way, the SI by itself cannot assess the real effects of the restrictions on pandemic transmission and, thus, cannot quantify the benefits in reducing the severe infection cases. For example, a certain country can implement the maximum level of restrictions (SI=100, full lockdown) without having the capacity to enforce them. Under those circumstances, the real SI value might have an offset because the citizens are partially following the imposed rules. This situation leads to unexpected results about the mitigation. Instead, an instance of a direct measure of the effects of the restrictions can be given by the daily number of infected people. But again, this latter variable is often an inaccurate metric due to several reasons, such as the impossibility to track many asymptomatic cases or, in particular for the first wave, the reduced ability in testing the population, thus often leading to a report relied upon an underestimation of daily cases. Hence, following the indications in [14], we chose the hospitalized ICU patient number thanks to its robustness with respect to other variables.

The distribution of the number of ICU hospitalized patients over time can be considered to assess the effectiveness of the imposed restrictions that are quantified by the SI. Indeed, it can represent an empirical probability distribution function measuring the probability that a person can enter the ICU because of a disease caused by the SARS-CoV-2 virus. It is worth to be remarked that the distributions for all the countries are temporally aligned, thus having that the first value into the sample space coincides with the day when the first cases have been reported in each country, as shown in Fig 1. A further issue to be addressed is how to compare the SI (a number) with a probability distribution function. To this aim, we need to sum up it. Hence, we computed, for each country, some of the most common synthetic indexes, i.e., the *moments* of the distribution, to characterize its shape. Our guess is indeed that if we have a tangible effect, it can be measurable through the modification of the shape of the probability distribution function of the ICU cases. Having characterized a distribution’s central value (i.e., the day when the half of the cases happened during the first wave), one conventionally characterizes its “width” or “variability” around that value. Most common is the variance, or, the related (its square root) *standard deviation*. For a random variable vector *Y* made up of *N* scalar observations,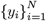, the sample standard deviation, *σ*_*Y*_, is defined as follows [16]:

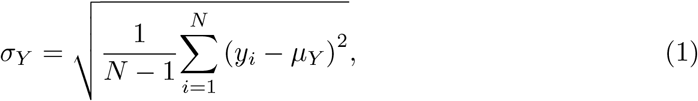

where *µ*_*Y*_ is the mean of *X*, i.e.:

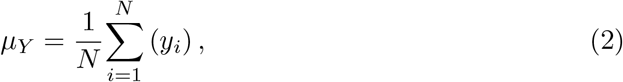

**Fig 1.**
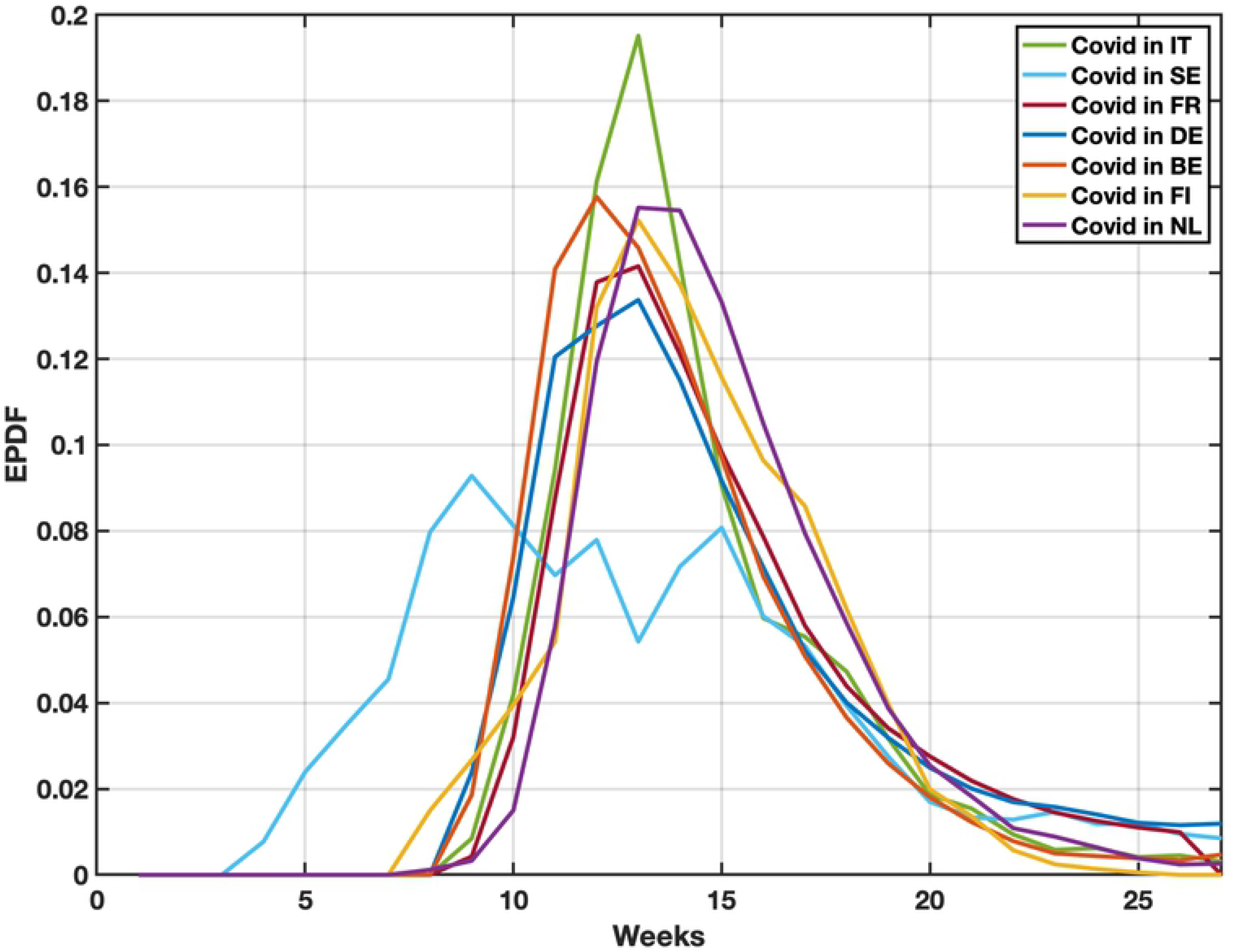
ICU weekly patient number temporal distribution. ICU weekly patient number temporal distribution for the seven analyzed western European countries normalized and lined up.

The higher the variance (or, equivalently, the standard deviation), the greater the variability of the distribution around its mean.

The *skewness* instead characterizes the degree of asymmetry of a distribution around its mean. While the standard deviation is non-dimensional quantity, that is, have the same units as the measured quantities, the skewness is a pure number that characterizes only the shape of the distribution. It is defined for the random variable *Y* as follows [16]:

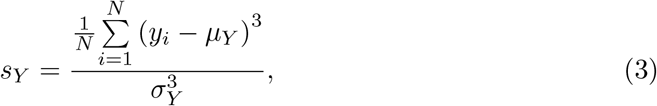

where *s*_*Y*_ is the skewness on the *N* scalar observations, 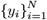, of *Y*. For a unimodal distribution, negative skew indicates that the tail is on the left side of the distribution, and positive skew the vice versa. A zero value means that the tails on both sides of the mean balance out overall. The normal distribution has a skewness equal to 0 (that is symmetric).

The *kurtosis* is also a non-dimensional quantity. It measures the relative peakedness or flatness of a distribution. The index is defined for the random variable *Y* as follows [16]:

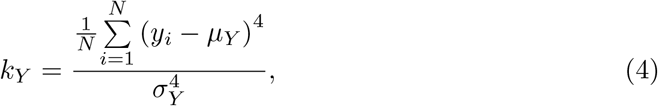

where *k*_*Y*_ is the kurtosis on the *N* scalar observations, 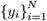, of *Y*. A distribution with a kurtosis greater than 3 is termed *leptokurtic*, less than 3 is termed *platykurtic*, and equal to 3 is called *mesokurtic* (i.e., the kurtosis of the normal distribution). Leptokurtic distribution has heavier (fatter) tails than the normal distribution. Instead, platykurtic distribution has tails less heavy than the ones of the normal distribution.

Afterwards, we assess the correlation among the previously cited statistical indicators and the SI averaged during the first wave. Given 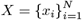 representing the set of *N* SIs, where *N* indicates the total number of the analyzed countries, and 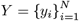 the related set of each synthetic index (either standard deviation or skewness or kurtosis), we exploit the *multivariate regression framework* pointing out the possible linear correlation between *X* and *Y*. Furthermore, Pearson’s correlation coefficient, *ρ*_*X,Y*_, is computed to measure the linear correlation between *X* and *Y*. It is defined as follows:

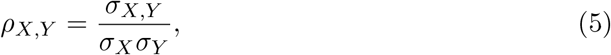

where *σ*_*X*_ and *σ*_*Y*_ are the two standard deviations for *X* and *Y* calculated as in (1), and *σ*_*X,Y*_ is the sample covariance:

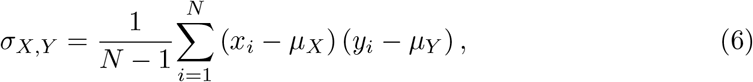

with *µ*_*X*_ and *µ*_*Y*_ computed as in (2). *ρ*, is a normalized measurement of the covariance, such that the result always has a value between −1 and 1. It is worth to be noted that the measure can only reflect a linear correlation of variables ignoring other types of relationships. A value of 1 implies that exist a linear equation describing the relationship between *X* and *Y*, with all data points lying on a line. The correlation sign is determined by the regression slope: a value of +1 implies that all data points lie on a line for which *Y* increases as *X* increases (i.e., *X* and *Y* are perfectly positively correlated), and vice versa for −1 (i.e., *X* and *Y* are perfectly negatively correlated). A value of 0 implies that there is no linear dependency between the two variables.

After selecting those indicators showing a strong correlation with the SI, we assess the significance level reached by the selected indicators. Namely, even if we find that the skewness can be used as a proxy to measure the efficiency of the imposed restrictions, we do not know which is the threshold upper that this index indicates a relevant effectiveness of the measurements (i.e., sensitively greater than the no restriction case). To deal with this issue, we take those indicators showing a strong correlation with SI and we assess their consistency through the pre-pandemic flu outbreaks. These scenarios represent similar cases of spreading of respiratory viruses in a given country with no restriction. Under the hypothesis that SARS-CoV-2 impacts on the shape of the probability distribution of the ICU cases as the seasonal flu, we can measure if there is a clear divergence in the used indicators, computed on both pre-pandemic flu outbreaks and on the first wave of the outbreak for a given country. This is enough to state if the imposed restrictions provided an effect that is significantly different than that of the no restriction case. In our work we carried out the analysis for Italy using pre-pandemic flu outbreaks in the following years: 2014-15, 2016-17, 2017-18, and 2018-19.

## Results

Before computing the statistical analysis, the empirical distribution probability distribution function of the ICU hospitalizations for each country is shown in Fig. 1.

In Tab.2 we report the values of the main statistical indicators, i.e. skewness, kurtosis and standard deviation, and the Stringency Index for the countries under investigation.

**Table 2.** ICU statistical parameters and SI. ICU statistical parameters and Stringency Index for the seven countries under study.

| Country | Skewness | Kurtosis | Std. Dev. | SI |
| --- | --- | --- | --- | --- |
| Belgium | 1.29 | 5.14 | 3.22 | 52.40 |
| Finland | 0.30 | 2.92 | 2.89 | 41.45 |
| France | 1.17 | 3.70 | 4.34 | 58.02 |
| Germany | 1.12 | 3.87 | 4.00 | 59.14 |
| Italy | 1.34 | 5.20 | 3.25 | 67.06 |
| Sweden | 0.62 | 3.03 | 4.92 | 49.40 |
| The Netherlands | 0.93 | 4.16 | 2.96 | 53.88 |

Figures 2, 3 and 4 show the multivariate regression of the statistical parameters computed through Eqs.1, 3 ans 4 versus the Stringency Index. For the standard deviation, no significant correlation is found (*R*^2^=0.10).

**Fig 2.**
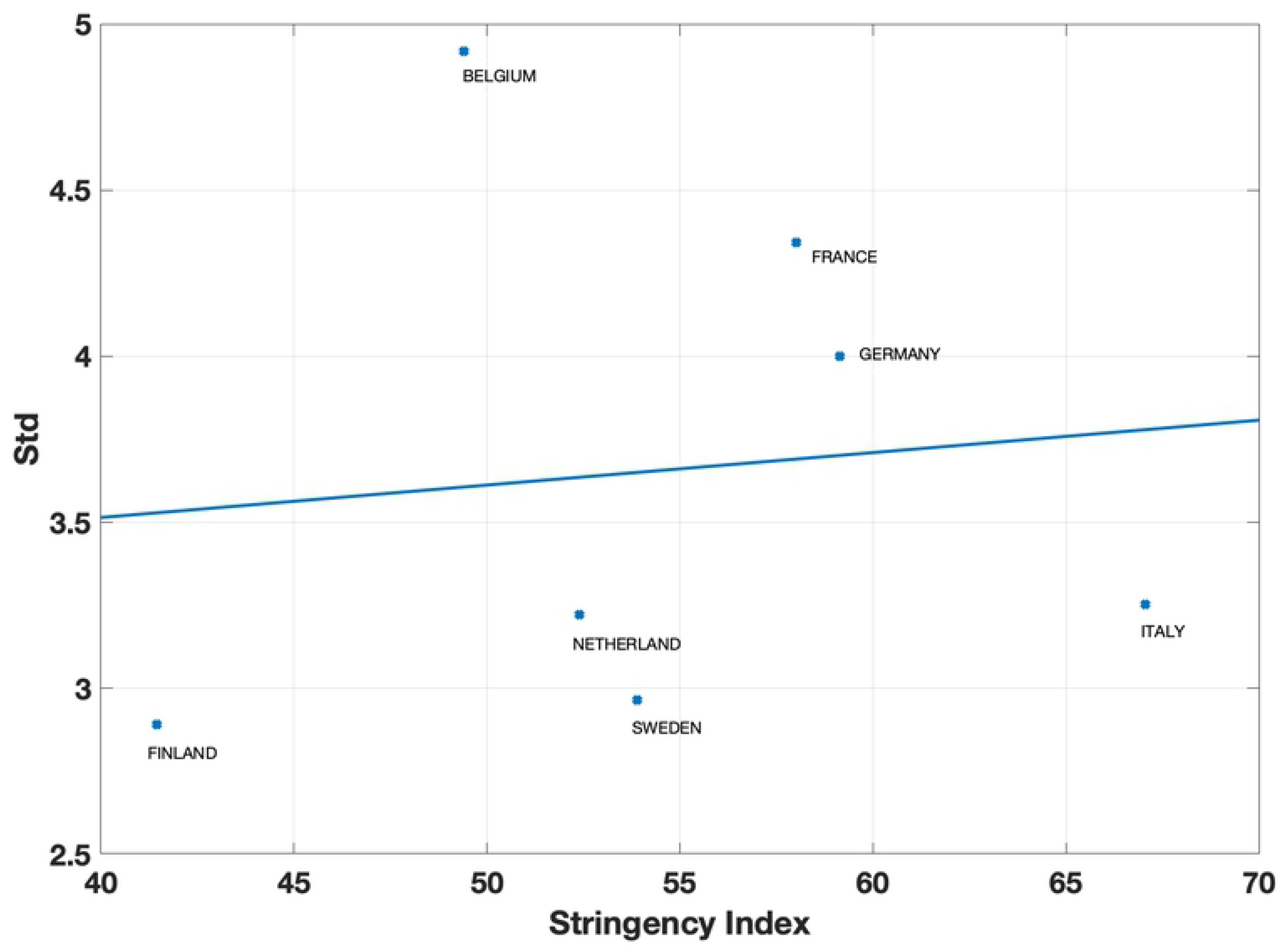
Correlation Std. Vs. SI. The multivariate regression between the Stringency Index and the Standard Deviation

**Fig 3.**
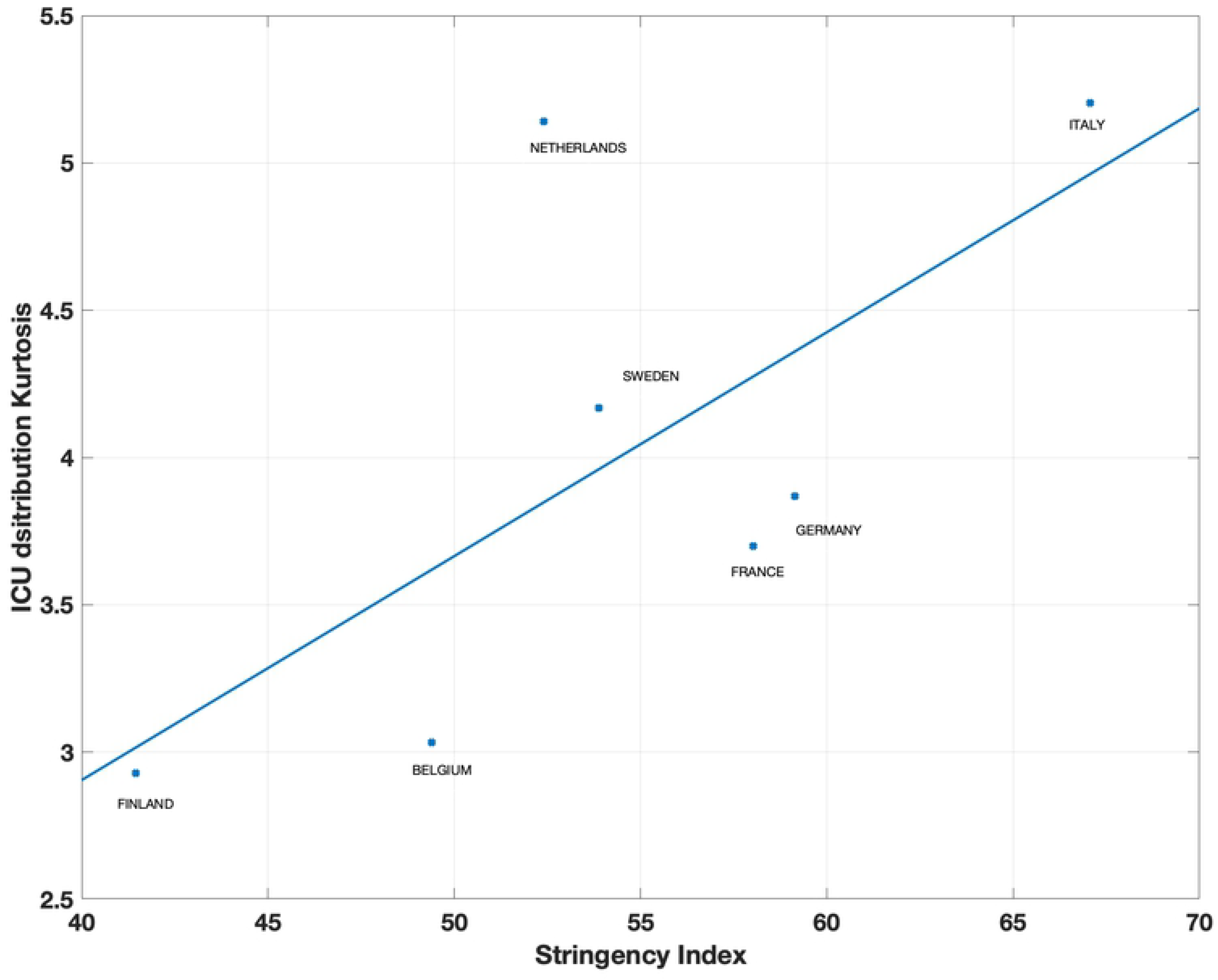
Correlation Kurtosis. Vs. SI. The multivariate regression between the Stringency Index and the Kurtosis

**Fig 4.**
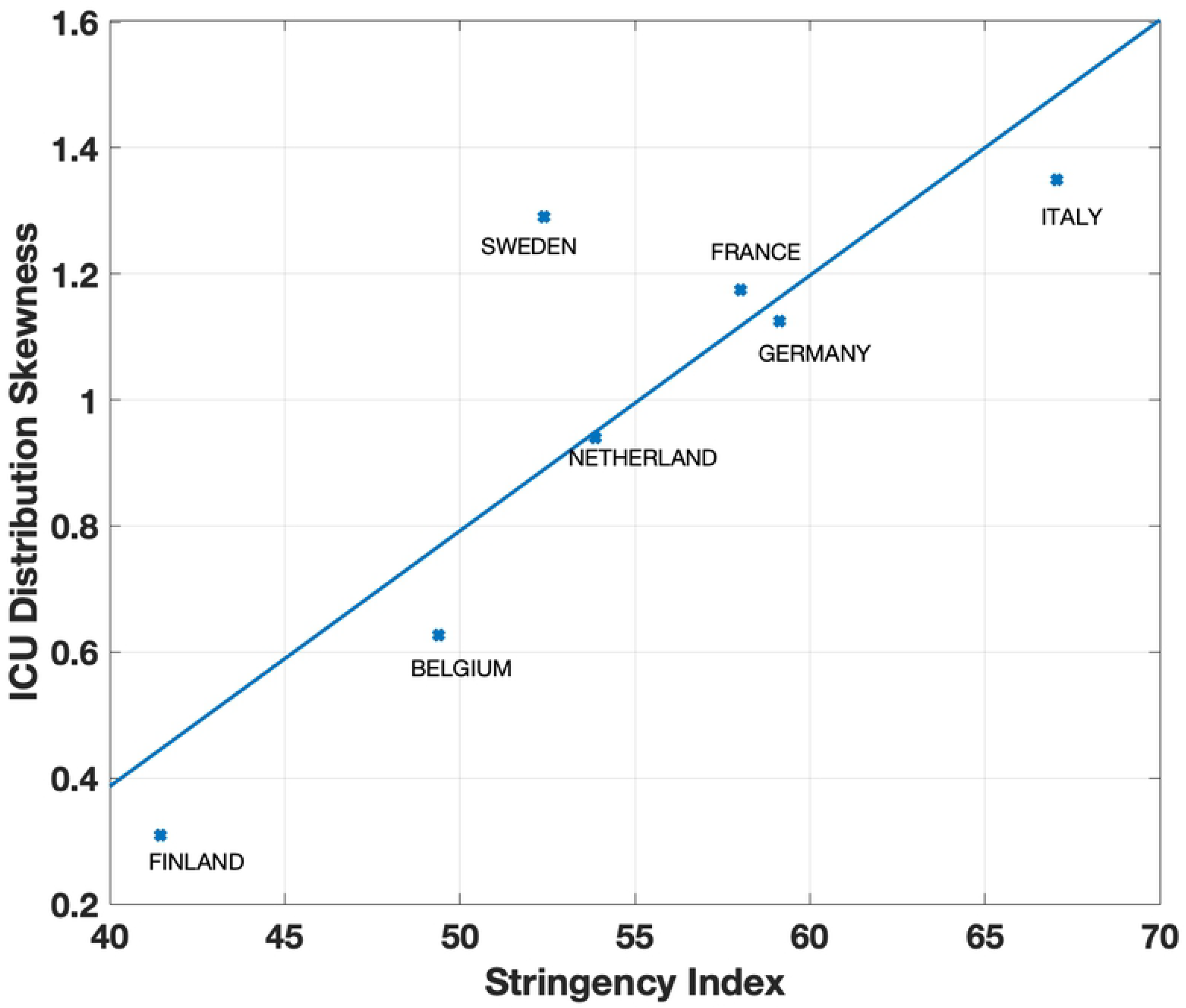
Correlation Skewness. Vs. SI. The multivariate regression between the Stringency Index and the Kurtosis

Instead, the multivariate regression between the kurtosis and the Stringency Index shows a certain degree of correlation, but still weak (*R*^2^=0.67).

The skewness instead, refers to a distortion or asymmetry that deviates from the perfect bell-shaped distribution.

In this case, the multivariate regression of the skewness and the Stringency Index is very high, with an *R*^2^=0.87. This result implies that the Stringency Index accounts almost for 90% of the variability of the skewness. For this reason, this distribution moment is the most appropriate to the evaluate the effects of the restrictions on the pandemic outbreak. But, as previously stated, a correlation with the SI doesn’t prove that the statistical index can be considered a valid proxy for the implemented restrictions.

To test the consistency of the skewness, we compute the values of this moment of the ICU distribution for the non-pandemic flu outbreaks during 2014-15, 2016-17, 2017-18, 2018-19 in Italy where no restrictions were applied, as shown in Figure 5

**Fig 5.**
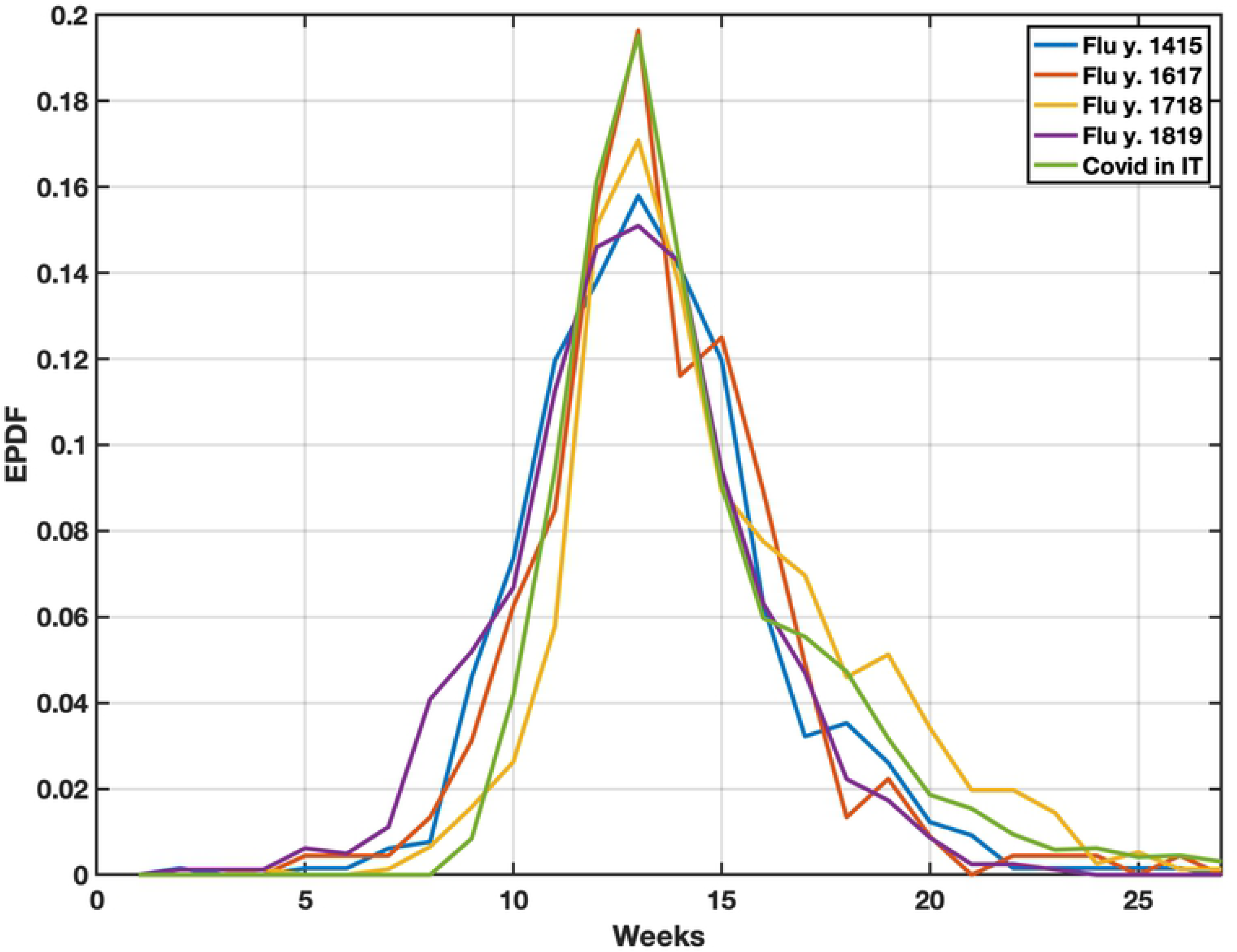
ICU distribution for Italy normal flu and COVID-19. ICU distributions for normal flu(2014-15, 2016-17, 2017-18, 2018-19) and COVID-19 outbreak

The results are reported in Tab.3

**Table 3.** Skewness for ICU distribution for normal flu and COVID-19 outbreaks. COVID-19 Skewness is highlighted as show a much higher value with respect to the normal flu values Skewness for normal flu outbreak and COVID-19 pandemic outbreak.

| Year | Skewness |
| --- | --- |
| 2014-15 | 0.53 |
| 2016-17 | 0.58 |
| 2017-18 | 0.77 |
| 2018-19 | 0.39 |
| COVID-19 2020 | **1.35** |

The results highlight that the skewness values for the non pandemic flu outbreaks are substantially different from the skewness value for the COVID-19 pandemic outbreak. This finding confirms that the skewness is a significant variable that can be used as a proxy to measure the effectiveness of the implemented restrictions at government level.

## Discussion and Conclusions

We assessed the restrictions effectiveness, implemented at government level by seven western European countries to curb the recent COVID-19 pandemic outbreak. The analysis is carried on through a statistical analysis. We considered, as a reliable variable to represent the COVID-19 pandemic outbreak, the weekly number of hospitalized patients into the Intensive Care Unit (ICU). This variable has already be proven effective in some previous analysis on COVID-19 pandemic outbreak [14]. The different countries ICU distributions are normalized to the maximum value and temporally shifted to make the maximum coincident (Fig. 1). The implemented restrictions can be represented by a synthetic index, the Stringency Index (SI; [15]), which ranges from 0 (no restrictions) to 100 (full lockdown). The first step of the analysis consists in testing the correlation of the main statistical parameters characterizing the ICU distribution, e.g. the skwewness, the kurtosis and the standard deviation with respect to the averaged SI value over the first COVID-19 pandemic outbreak (January-May 2020). A significant correlation is found with the skewness (*R*^2^=0.87), while standard deviation and kurtosis poorly (*R*^2^=0.10) and moderately correlate (*R*^2^=0.67). As further step, we compared for Italy the skewness value during COVID-19 with those obtained for normal flu outbreaks prior to 2020. The intercomparison permits to evaluate if the skewness is a significant parameter effectively linked to the implemented restrictions. The analysis show that for the normal flu the skewness is much lower, as much as 70% with respect to the first-wave COVID-19 outbreak where social restrictions were applied. This proofs, at least for Italy, assuming that the COVID-19 and normal flu outbreaks have similar behaviours that the implemented restrictions impact the ICU temporal distribution modifying the skewness. As a final statement, we can say that the restrictions produced a measurable effect on the ICU distribution, but it is impossible to estimate the effects in terms of infection reduction and saved lives. As stated above, a limitation of this study is given by the fact that we assume that the normal flu and the COVID-19 pandemic outbreak have the same trend. As a future perspective, it should be investigated the possibility to link the skewness to the number of ICU hospitalized patients, in order to assess the restriction effectiveness in saving lives.

## Data Availability

All dataset is freely available online https://www.bsg.ox.ac.uk/research/research-projects/covid-19-government-response-tracker

